# Lower Hippocampal Volume Partly Mediates the Association Between rs6859 in the *NECTIN2* Gene and Alzheimer’s Disease: New Findings from Causal Mediation Analysis of ADNI Data

**DOI:** 10.1101/2025.09.02.25334930

**Authors:** Aravind Lathika Rajendrakumar, Konstantin G. Arbeev, Olivia Bagley, Anatoliy I. Yashin, Svetlana Ukraintseva, the Alzheimer’s Disease Neuroimaging Initiative

## Abstract

Infections may contribute to neurodegeneration, including Alzheimer’s disease (AD). Polymorphism in the *NECTIN2* gene has been linked to both AD and vulnerability to infections. We hypothesized that neurodegeneration may mediate the connection between this polymorphism and AD. To test this hypothesis, we conducted a causal mediation analysis (CMA) using the Alzheimer’s Disease Neuroimaging Initiative (ADNI) data. We found that smaller hippocampal volume (HV), a biomarker of neurodegeneration, significantly mediated the association between rs6859 in *NECTIN2* and AD risk. For the right HV, the mediated effect was 42.75%, while for the left HV, it was 49.76%. In linear mixed models (LMM), carrying the rs6859 risk alleles (A) was associated with a reduction in right HV (β = -0.16, *p* = 0.03), left HV (β = -0.14, *p* = 0.04), and total HV (β = -0.15, *p* = 0.04). In this data, the rs6859 (A) was a risk factor for AD only in men. Our results suggest that hippocampal atrophy may substantially mediate the association between *NECTIN2* polymorphism and AD risk.

## Introduction

Alzheimer’s disease (AD) is a major cause of neurodegeneration and cognitive impairment in older adults.^1^ Hippocampal atrophy is a key indicator of neurodegeneration and serves as a major biomarker of AD pathology. Hippocampal volume measured by magnetic resonance imaging (MRI) may detect pathological changes that are useful for predicting dementia even in the absence of clinical symptoms.^2^ Progressive atrophy of brain structures occurs in AD due to neurodegenerative changes that worsen with the severity of the disease.^3^ The hippocampus is the part of the brain responsible for memory formation.^4^ Pathological features of AD typically first manifest in and around this region.^5^

AD is a multifactorial disorder arising from the interplay of various factors, including genetic variation and infections, among others.^6–13^ An increasing body of research suggests that infections may play an important role in AD, dementia, and neurodegeneration.^9,12,14–21^ Seemingly mild infections, such as urinary tract infections (UTIs), can profoundly elevate inflammation levels^22^, and disrupt hippocampal nerve plasticity.^23^ These changes are difficult to reverse even with appropriate medications.^23^ Infections can also impair hippocampal metabolism.^24^ Despite this, it is not yet clear whether the infections are just a triggering factor or a direct cause of AD.^8^ Most of the evidence comes from observational studies, many of which may be subject to bias. Employing causal mediation analysis (CMA) with genetic risk factors involved in both AD and infections may provide better insights into the processes underlying AD development.

Genetic variation in the *NECTIN2* gene is a plausible candidate for such analysis. It has been associated with both AD and vulnerability to infections, especially to herpes viruses.^10,25^ The *NECTIN2* product participates in the maintenance of cellular tight junctions and neurons.^26,27^ Hence, a variation in this gene could potentially influence pathogenic spread in the brain.^27,28^ A single nucleotide polymorphism (SNP) rs6859 in *NECTIN2* is one of the strongest AD risk factors confirmed by GWAS.^10^ It has been associated with cognitive changes, phosphorylated tau, pneumonia, and shingles vaccine efficacy against AD.^11,12,29,30^ We recently found that prior infections and the rs6859 risk allele (A) are associated with reduced hippocampal volume in the UK Biobank participants.^20^ This indicates a possibility that hippocampal atrophy might be one of the mediators of the detrimental effect of rs6859 (A) on AD risk.

Here, we investigate whether the rs6859 (A) allele, a risk factor for AD involved in vulnerability to infections, influences hippocampal volume in ADNI data. We also employ causal mediation analysis to examine whether hippocampal atrophy, reflected in lower hippocampal volume, mediates the association between rs6859 (A) in *NECTIN2* and AD risk.

## Materials and Methods

### Study Population, Hippocampal Volume Measurement, and Genetic Data Extraction

Data used in the preparation of this article were obtained from the Alzheimer’s Disease Neuroimaging Initiative (ADNI) database (adni.loni.usc.edu). The ADNI was launched in 2003 as a public-private partnership, led by Principal Investigator Michael W. Weiner, MD. The primary goal of ADNI has been to test whether serial magnetic resonance imaging (MRI), positron emission tomography (PET), other biological markers, and clinical and neuropsychological assessment can be combined to measure the progression of mild cognitive impairment (MCI) and early AD. We performed a secondary data analysis, and no participants were directly enrolled in our study. In ADNI, consenting participants are enrolled through a staggered recruitment approach and have varying follow-up times.

Medical history, vital signs, and other clinical parameters were collected at screening. Neuroimages and biomarkers were measured in a subset of participants based on a standard protocol developed by clinical imaging experts.^31^ Hippocampal volumetric data in ADNI were acquired using 1.5 Tesla (T) and 3 Tesla (T) MRI scanners from selected manufacturers and were automatically extracted using FreeSurfer software.^31–33^ Genotyping in the ADNI was conducted using different genotyping arrays: Human610-Quad BeadChip, Illumina HumanOmniExpress BeadChip, and the Illumina Infinium Global Screening Array v2 (GSA2), and was stored in the Plink format.

### Data linkage

We linked the hippocampal volume data with demographics, SNP rs6859, and clinical variables, including the use of diabetes medications by their participant roster ID (RID) (http://adni.loni.usc.edu). Diabetes medication use (Yes/No) was identified from the prescription data by matching the drug names present in the Anatomical Therapeutic Chemical (ATC) classification system coding (https://www.who.int/tools/atc-ddd-toolkit/atc-classification). We relabeled the smoking and alcohol history as Ever/Never and extracted the SNP rs6859 alleles using the --recode command from the Plink 1.90 beta version.^34^ These were further verified with the summary SNP information file to ensure accurate data linkage.

### Statistical Analysis

All statistical analyses were performed using R software version 4.3.2.^35^ We included age, hippocampal volumes, diabetes (yes/no), SNP rs6859, smoking (ever/never), alcohol use (ever/never), visits, duration of education, race, and married status (ever/never) in the regression models. Continuous variables were summarized as means ± standard deviation, and categorical variables were represented as frequencies and percentages. Univariate and multivariate visualizations were generated with the *ggplot2* package.^36^ Histograms of hippocampal volume and its longitudinal trajectories by age and clinical visits, stratified by rs6859 allele status, are presented in the Supplementary Material. We applied an ordered quantile normalization (ORQ) transformation^37^ using the *bestNormalize* package for data normalization of hippocampal volumes.^38^ This kind of transformation is suitable for improving the fit of parametric models. First, adjusted linear mixed models using the *lme4* package were run to assess the association of SNP rs6859 with normalized hippocampal volumes over time.^39^ Additionally, total hippocampal volume (sum of right and left hippocampal volumes) was considered an outcome.

Allele dosages of SNP rs6859 were included in the linear and logistic regression models for the mediator and outcome, assuming an additive genetic model. Age, sex, marital status, number of visits, education duration, diabetes, race, smoking, and alcohol use were controlled in the analysis. The “*dredge*” function in the *MuMin* package was used for variable selection.^40^ It allows a reproducible and automated model selection by ensuring that all possible combinations of model terms are considered for determining the parsimonious model based on the Akaike Information Criterion (AIC). A random intercept was specified for different starting values of hippocampal volumes and random slopes for differences in clinical visits across individuals. The data were apportioned accordingly for sex-stratified analysis, and associations were computed for these groups. Statistical associations for the regression models were considered significant for a two-sided p-value less than 0.05.

We next estimated the parameters from the causal mediation analyses (CMA) using median values from all readings for hippocampal volumes and other continuous covariates generated by the *dplyr* package.^41^ Given the extreme values in the predictors, a more stable measure could be achieved compared to using the average readings.^42^ CMA mimics different scenarios by varying the exposure relationship with the outcome, conditional on the mediator values likely at different levels of exposure to influence the potential outcomes.^43^ As we have a continuous mediator and a binary outcome (AD), the CMA method can be used to reliably decompose the exposure effects into natural direct and indirect effects.^44^ To investigate if the *NECTIN2* gene polymorphism influences AD risk through hippocampal volume reduction, we conducted a covariate-adjusted mediation analysis using the *Medflex* package.^45^ We used the *RNomni* package to transform the summarized hippocampal measures.^46^ The main advantages of *RNomni* are that it applies Rank-based inverse normal transformation (INT), which is particularly useful for small sample sizes, and variability in the model is ensured while estimating associations. Only those predictors of AD selected by the algorithm were carried forward to the CMA for covariate adjustment. We used the “neWeight” function to apply the ratio-of-mediator-probability weighting (RMPW) method in the same package to create a pseudo dataset for counterfactual estimation, using inverse probability weighting.^45,47^ At first, an expected value for the mediator under observed and counterfactual levels of exposure was computed. These estimated values were used by the neWeight() function to create an in-built pseudo dataset by reweighting on the expected mediator value computed earlier. In this way, subject-specific weights are calculated, which are then subsequently accounted for in a regression that models the exposure-mediator-outcome relationship and finally provides separate coefficients of the Natural Direct Effect (NDE) and Natural Indirect Effect (NIE).^45,48^ Due to the logit scale, we have also interpreted the NDE and NIE as odds ratios for better understanding. We computed the proportion of mediated effects (PE) by dividing NIE by Total effects (TE), since the package lacks a built-in function for this calculation.

## Results

### Sample Characteristics

A total of 902 records belonging to 318 participants were analyzed, as detailed in the flow chart (Supplementary Figure S1). The histograms in Supplementary Figure S2 show that the right hippocampal volume had a slightly leptokurtic distribution in comparison to the left hippocampal volume. There were differences in the trajectories of hippocampal volumes with age when stratified by rs6859 allele status, as depicted in Supplementary Figure S3. For the younger age range, the hippocampal volume trajectories remained stable. Among the participants, those with the GG genotype had a rapid decline with age for the right hippocampal volume. With aging, participants in the GA group experienced a faster decline in LHV compared to those in the GG group, showing a different trend from that observed in RHV. The change in hippocampal volumes for the AA genotype was not profound for either hippocampal region, or the relative difference between the age groups visualized was minimal. Longitudinal changes in hippocampal volume across clinical visits, stratified by rs6859 allele status, are shown in Supplementary Figure S4. Carriers of the rs6859 A risk allele exhibited decreased hippocampal volumes with an increasing number of clinical visits.

### Predictors of right hippocampal volume and differences by sex

Table 1 shows the coefficients of variables associated with the longitudinal variations in right hippocampal volume over time. Increased age and clinical visits resulted in lower hippocampal volume (*p*<0.001). Being female has a greater effect on right hippocampal volume reduction than having diabetes. Carriage of A alleles of rs6859 predicted a lower right hippocampal volume than non-carriers (-0.155, *p*= 0.033). Marital status, although included in the model, did not achieve statistical significance. AIC-based regression identified a similar set of variables for RHV in both sexes (Supplementary Tables S1 and S2). Notably, the variable selection methods chose age and behavioral risk factors, such as alcoholism, which were not identified for LHV or in the subgroups. However, their influences differed, showing more prominence in males. None of the variables, excluding age and sex, were predictive in females. Across both sexes, SNP rs6859 was not significant, although the estimate was larger and approached the significance threshold (*p* = 0.06) in males. Similarly, as seen in the case of LHV, males with diabetes were more likely to have a lower RHV.

**Table 1.** Linear mixed model estimates of rs6859 and covariates with right hippocampal volume (n=318, observations=902)

| Variables | Estimates | 95% CI | *p* |
| --- | --- | --- | --- |
| Age | -0.062 | -0.074, -0.050 | 0.000*** |
| Diabetes (Yes) | -0.376 | -0.637, -0.115 | 0.005** |
| Married Status (Never) | 0.365 | -0.069, 0.799 | 0.102 |
| rs6859 | -0.155 | -0.297, -0.013 | 0.033* |
| Sex (Female) | -0.630 | -0.832, -0.427 | 0.000*** |
| Visits | -0.022 | -0.033, -0.012 | 0.000*** |

**Note.** Estimates adjusted for education, race, smoking, alcohol, and diabetes. Abbreviations: CI, Confidence Interval; *p*, *p-value*.
\* $p < 0.05$ ; \*\* $p < 0.01$ ; \*\*\* $p < 0.001$ .

## Predictors of left hippocampal volume and differences by sex

Table 2 presents the results of the LMM analysis for the left hippocampal volume. In the whole dataset, an increase in the dosage of the rs6859 A allele was associated with a reduction in left hippocampal volume over time (β = -0.139, p=0.044). Females had an increased risk of hippocampal volume loss (β = -0.584, p<0.001). Increasing age, clinical visits, and a diagnosis of diabetes were also identified as risk factors influencing left hippocampal atrophy. No significant influence of education or marital status on the evolution of left hippocampal volume was observed. Supplementary Tables S3 and S4 display the results of the sex-stratified analysis. All the variables included in the final model for males adversely affected LHV and were statistically significant. Most importantly, SNP rs6859 was selected (-0.242, p=0.014) only in this category. In contrast, a different set of variables was chosen in females, except for age and clinical visits, which had the same direction of effect as in males. Higher education and never being married were associated with improved LHV, with the former demonstrating a borderline significance (p = 0.05).

**Table 2.** Linear mixed model estimates of rs6859 and covariates with left hippocampal volume (n=318, observations=902)

| Variables | Estimates | 95% CI | *p* |
| --- | --- | --- | --- |
| Age | -0.058 | -0.070, -0.047 | 0.000*** |
| Diabetes (Yes) | -0.284 | -0.539, -0.044 | 0.025* |
| Education | 0.032 | -0.005, 0.071 | 0.093 |
| Married Status (Never) | 0.360 | -0.079, 0.748 | 0.08 |
| rs6859 | -0.139 | -0.269, 0.003 | 0.044* |
| Sex (Female) | -0.584 | -0.782, -0.383 | 0.000*** |
| Visits | -0.025 | -0.0343, -0.015 | 0.000*** |

**Note.** Estimates adjusted for race, smoking, and alcohol. Abbreviations: CI, Confidence Interval; *p*, *p-value*.
\* $p<0.05$ ; \*\* $p<0.01$ ; \*\*\* $p<0.001$ .

### Predictors of total hippocampal volume and differences by sex

Table 3 details the variables associated with total hippocampal volume. The results showed a similar pattern to that estimated for LHV. Compared to LHV, the mixed model estimate for SNP rs6859 on THV was marginally higher (-0.145, 95% CI: -0.285, -0.004, p = 0.043) but lower than RHV. Age, sex, diabetes, and clinical visits were strongly associated with total hippocampal volume loss. Education and marital status variables were retained but were non-significant in the final model.

**Table 3.**
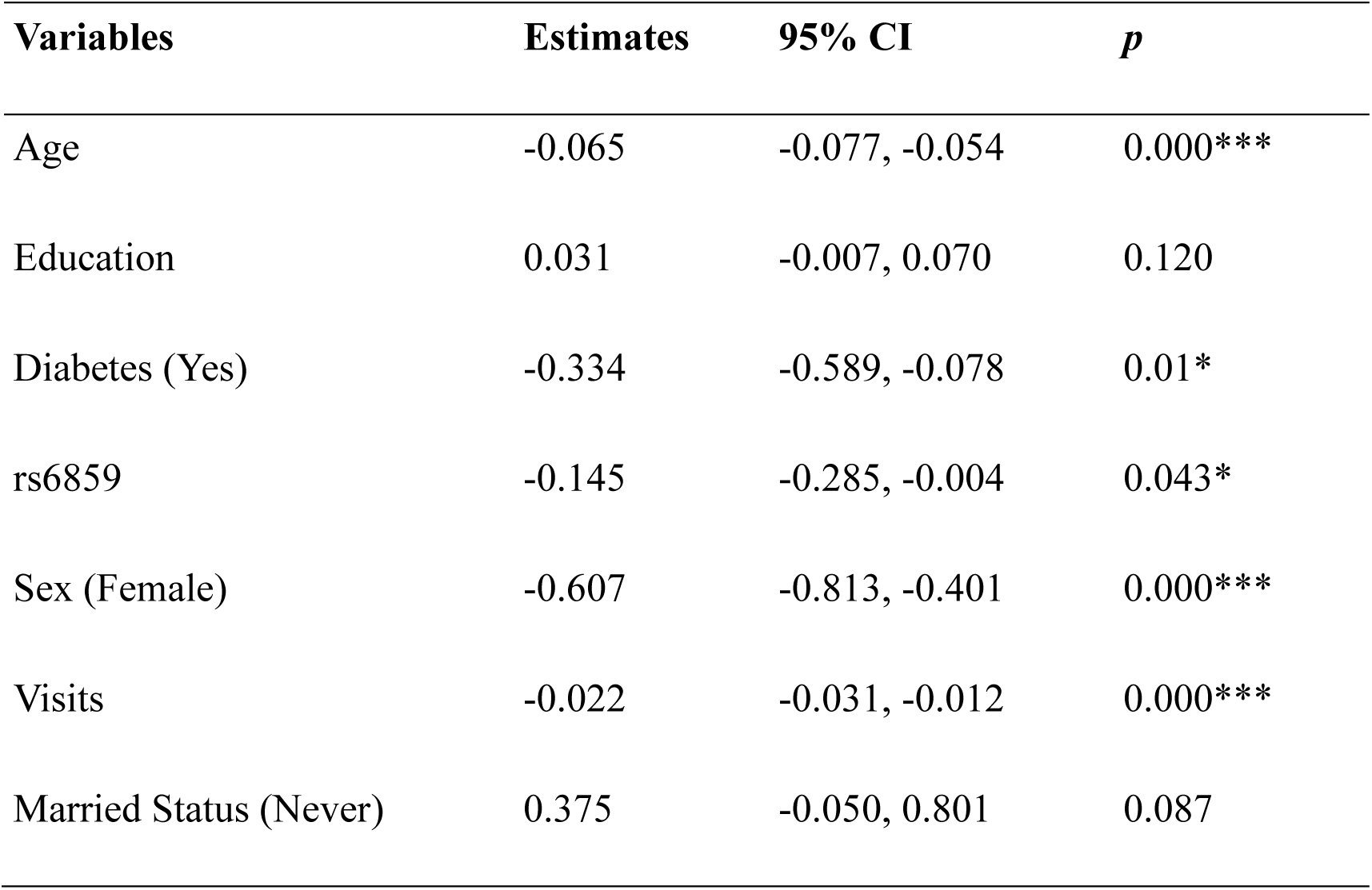
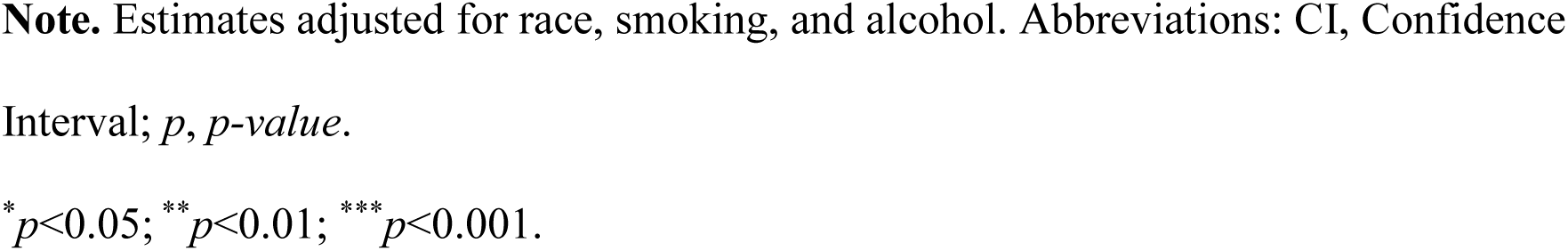
Multivariate response regression estimates for rs6859 and covariates for THV.

| Variables | Estimates | 95% CI | *p* |
| --- | --- | --- | --- |
| Age | -0.065 | -0.077, -0.054 | 0.000*** |
| Education | 0.031 | -0.007, 0.070 | 0.120 |
| Diabetes (Yes) | -0.334 | -0.589, -0.078 | 0.01* |
| rs6859 | -0.145 | -0.285, -0.004 | 0.043* |
| Sex (Female) | -0.607 | -0.813, -0.401 | 0.000*** |
| Visits | -0.022 | -0.031, -0.012 | 0.000*** |
| Married Status (Never) | 0.375 | -0.050, 0.801 | 0.087 |

**Note.** Estimates adjusted for race, smoking, and alcohol. Abbreviations: CI, Confidence Interval; $p$ , $p$ -value.
\* $p < 0.05$ ; \*\* $p < 0.01$ ; \*\*\* $p < 0.001$ .

Regarding the sex-stratified analysis, more variables were included for males (Supplementary Tables S5 and S6). SNP rs6859, diabetes, age, and clinical visits were associated with THV in males. The smoking and alcohol history was also chosen among the predictors but was not significant. The estimated coefficient of THV showed that the only common variables between males and females were age and visits.

### Participant characteristics in the summarized mediation analysis

Table 4 gives the characteristics of participants included in the dataset for the mediation analysis. Most participants were older than 60 years, males, better educated, and of the white race. Few individuals had up to five clinical visits. Almost 22% reported having smoked during their lifetime. However, only a few participants reported ever consuming alcohol, which is unusual and may indicate a potential reporting problem. More than a quarter of the participants were diagnosed with AD, and about 18% had diabetes. Regarding hippocampal volume, the right hippocampal volume was slightly higher than the left. A large proportion of participants (∼74%) were carriers of the rs6859 risk allele (A), while about half of the participants carried the *APOE4* risk allele.

**Table 4.**
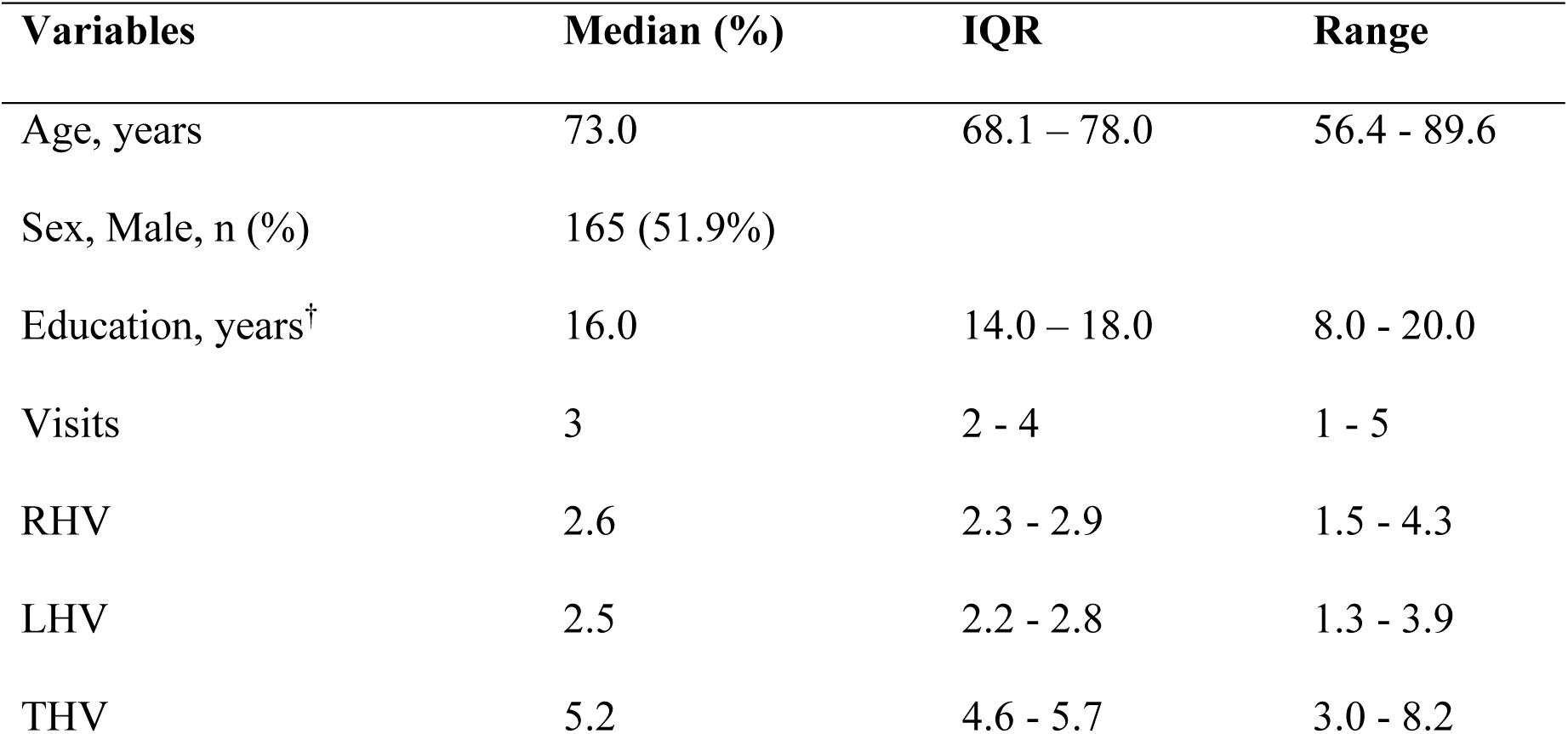
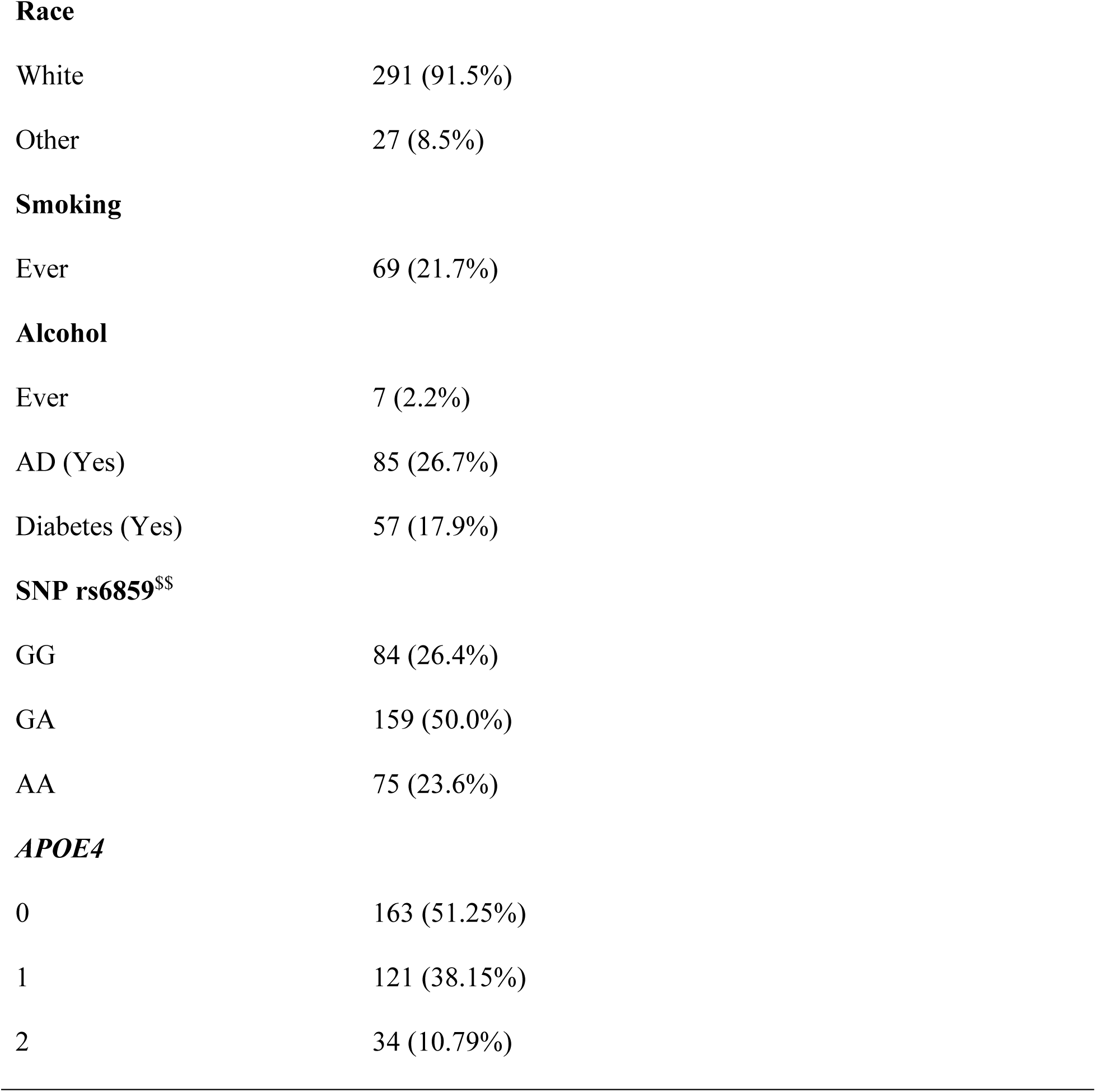
Participant characteristics in the mediation analysis sample.

| Variables | Median (%) | IQR | Range |
| --- | --- | --- | --- |
| Age, years | 73.0 | 68.1 – 78.0 | 56.4 - 89.6 |
| Sex, Male, n (%) | 165 (51.9%) |  |  |
| Education, years<sup>†</sup> | 16.0 | 14.0 – 18.0 | 8.0 - 20.0 |
| Visits | 3 | 2 - 4 | 1 - 5 |
| RHV | 2.6 | 2.3 - 2.9 | 1.5 - 4.3 |
| LHV | 2.5 | 2.2 - 2.8 | 1.3 - 3.9 |
| THV | 5.2 | 4.6 - 5.7 | 3.0 - 8.2 |

**Race**
|  |  |
| --- | --- |
| White | 291 (91.5%) |
| Other | 27 (8.5%) |

**Smoking**
|  |  |
| --- | --- |
| Ever | 69 (21.7%) |

**Alcohol**
|  |  |
| --- | --- |
| Ever | 7 (2.2%) |
| AD (Yes) | 85 (26.7%) |
| Diabetes (Yes) | 57 (17.9%) |

**SNP rs6859<sup>\$\$</sup>**
|  |  |
| --- | --- |
| GG | 84 (26.4%) |
| GA | 159 (50.0%) |
| AA | 75 (23.6%) |

***APOE4***
|  |  |
| --- | --- |
| 0 | 163 (51.25%) |
| 1 | 121 (38.15%) |
| 2 | 34 (10.79%) |

### Causal mediation analysis

Here, we quantified the direct and indirect effects of the A allele of rs6859 on the hippocampal volumes separately as well as jointly to gather evidence for a causal relationship. For this, we used only covariates deemed significant for AD and hippocampal volumes. The resulting estimates were interpreted as conditional probabilities of the mediator linked to the rs6859 and confounder levels in the model. First, we computed the estimates for the right hippocampal volume (Table 5), suggesting no direct effect of rs6859 on the right hippocampal volume (β = 0.220, *p* = 0.229). In contrast, we observed a mediating role for the RHV on the AD risk predicted by SNP rs6859 (β = 0.165, *p* = 0.033), implying a proportion of mediated effects (PME) of 42.75%. As expected, aging individuals were more susceptible to AD. Intriguingly, the number of visits was protective against AD risk in the model.

**Table 5.**
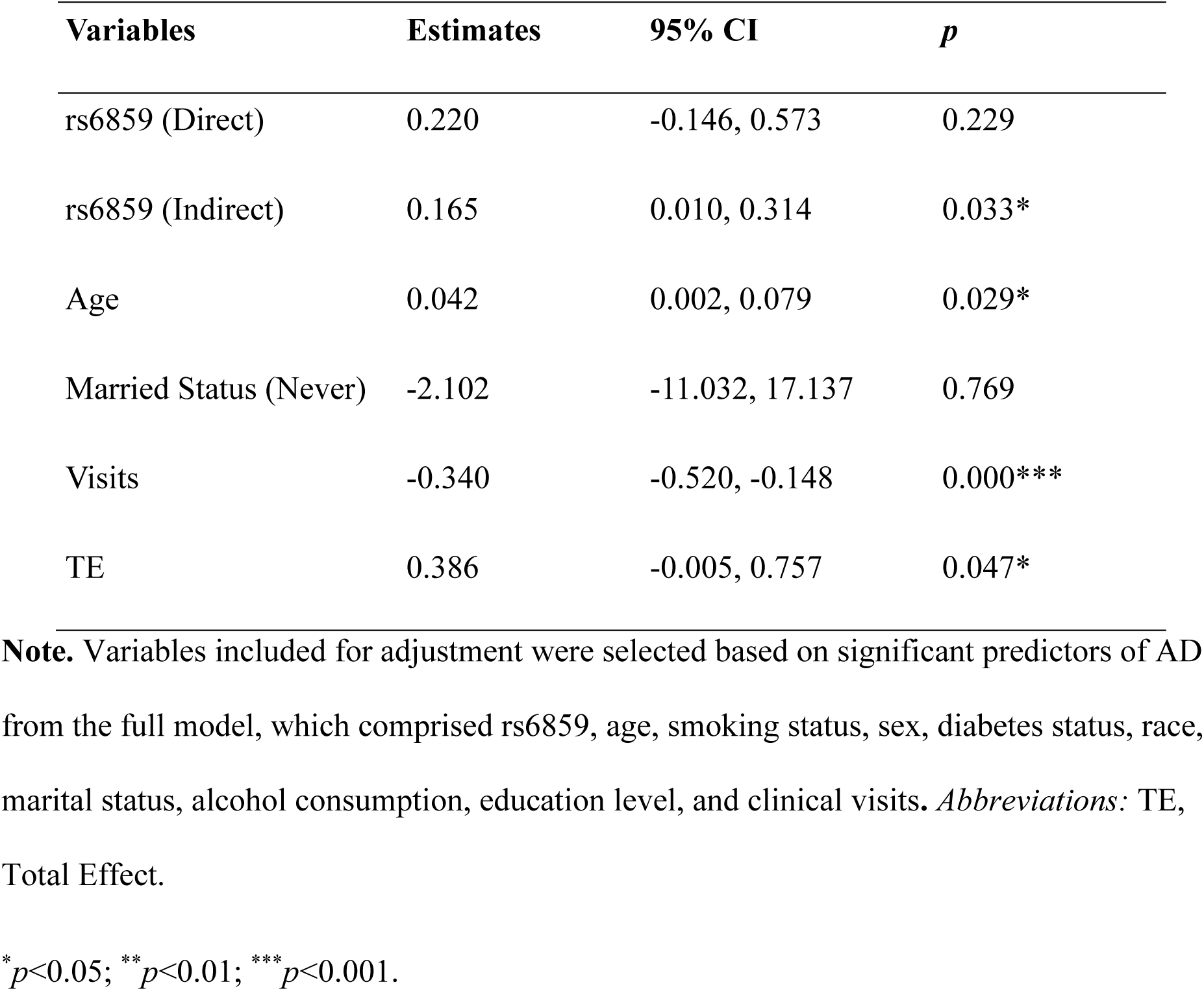
Confounder-adjusted mediation analysis showing the direct and indirect effects of rs6859 on AD through the right hippocampal volume.

| Variables | Estimates | 95% CI | *p* |
| --- | --- | --- | --- |
| rs6859 (Direct) | 0.220 | -0.146, 0.573 | 0.229 |
| rs6859 (Indirect) | 0.165 | 0.010, 0.314 | 0.033* |
| Age | 0.042 | 0.002, 0.079 | 0.029* |
| Married Status (Never) | -2.102 | -11.032, 17.137 | 0.769 |
| Visits | -0.340 | -0.520, -0.148 | 0.000*** |
| TE | 0.386 | -0.005, 0.757 | 0.047* |

**Note.** Variables included for adjustment were selected based on significant predictors of AD from the full model, which comprised rs6859, age, smoking status, sex, diabetes status, race, marital status, alcohol consumption, education level, and clinical visits. *Abbreviations:* TE, Total Effect.
\* $p < 0.05$ ; \*\* $p < 0.01$ ; \*\*\* $p < 0.001$ .

In Table 6, we provide the conditional probability estimates for the AD-rs6859 relationship, which is explained by left hippocampal volume change as a mediating variable. Once again, the direct effect for rs6859 was not statistically significant (p=0.259). We observed that the NIE observed was higher than that seen for the RHV, as reflected by a 4.80% increased risk. Adjusting for the covariate effects, LHV reduction was associated with a 22.7% higher risk for AD. Furthermore, the change in LHV with rs6859 risk alleles mediated a greater proportion of mediated effects, reaching 49.76%. A unit increase in age was associated with a 4.08% higher risk of AD. Contrary to expectations, a higher number of clinical visits offered protection against AD.

**Table 6.** Confounder-adjusted mediation analysis showing the direct and indirect effects of rs6859 on AD through the left hippocampal volume.

| Variables | Estimates | 95% CI | *p* |
| --- | --- | --- | --- |
| rs6859 (Direct) | 0.206 | -0.158, 0.559 | 0.259 |
| rs6859 (Indirect) | 0.205 | 0.039, 0.363 | 0.012* |
| Age (years) | 0.041 | 0.001, 0.078 | 0.035* |
| Married Status (Never) | -2.175 | -11.078, 16.995 | 0.761 |
| Visits | -0.341 | -0.521, -0.150 | 0.000*** |
| TE | 0.412 | 0.015, 0.789 | 0.036* |

**Note.** Variables included for adjustment were selected based on significant predictors of AD from the full model, which comprised rs6859, age, smoking status, sex, diabetes status, race, marital status, alcohol consumption, education level, and clinical visits. *Abbreviations:* Total Effect.
\* $p < 0.05$ ; \*\* $p < 0.01$ ; \*\*\* $p < 0.001$ .

Details of the causal mediation estimates for the direct and indirect effects of rs6859 on AD through total hippocampal volume (THV) are presented in Table 7. In the causal mediation analysis (CMA), there was no supporting evidence for a direct effect of rs6859 (estimate = 0.207, 95% CI: −0.157 to 0.558, *p* = 0.255) through changes in the mediator. However, the indirect effect of rs6859 was statistically significant (estimate = 0.190, 95% CI: 0.024 to 0.349, *p* = 0.021), indicating a 22.1% increased risk associated with each additional A allele, acting exclusively through hippocampal volume loss.

**Table 7.**
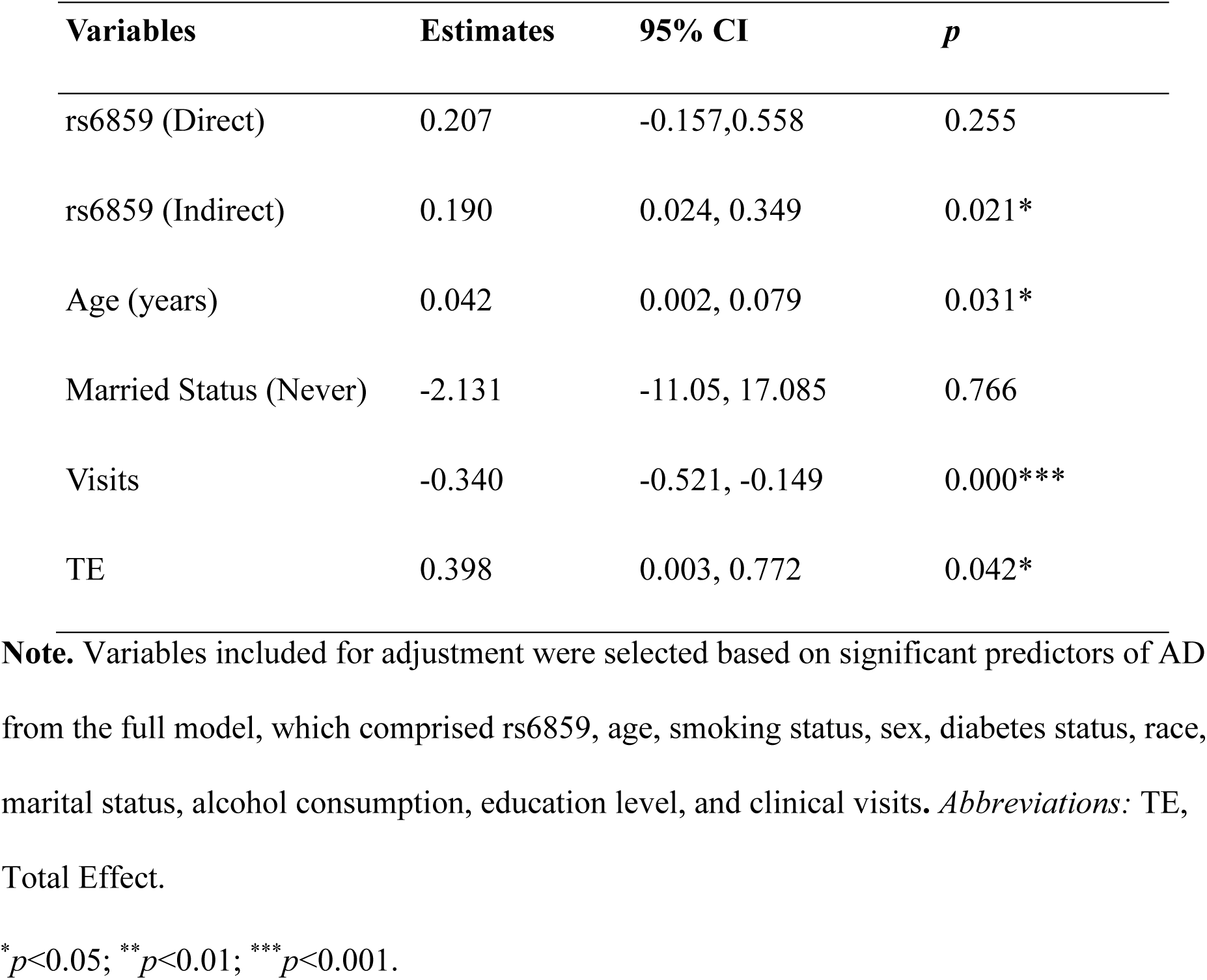
Confounder-adjusted mediation analysis showing the direct and indirect effects of rs6859 on AD through the total hippocampal volume.

| Variables | Estimates | 95% CI | $p$ |
| --- | --- | --- | --- |
| rs6859 (Direct) | 0.207 | -0.157, 0.558 | 0.255 |
| rs6859 (Indirect) | 0.190 | 0.024, 0.349 | 0.021* |
| Age (years) | 0.042 | 0.002, 0.079 | 0.031* |
| Married Status (Never) | -2.131 | -11.05, 17.085 | 0.766 |
| Visits | -0.340 | -0.521, -0.149 | 0.000*** |
| TE | 0.398 | 0.003, 0.772 | 0.042* |

**Note.** Variables included for adjustment were selected based on significant predictors of AD from the full model, which comprised rs6859, age, smoking status, sex, diabetes status, race, marital status, alcohol consumption, education level, and clinical visits. *Abbreviations:* TE, Total Effect.
\* $p<0.05$ ; \*\* $p<0.01$ ; \*\*\* $p<0.001$ .

Furthermore, the total effect (TE) was statistically significant, confirming that THV partially mediates the impact of rs6859 on AD. The computed proportion of mediated effect was 47.7%, suggesting that nearly half of the total effect is explained via the indirect pathway.

Additionally, covariate effects for THV remained consistent with those observed in other CMA analyses.

## Discussion

Our study found that the rs6859 (A) allele, a risk factor for AD in the *NECTIN2* gene involved in vulnerability to infections, is associated with reduced hippocampal volume in ADNI participants. This supports our recent finding in the UK Biobank.^20^

The CMA revealed that a lower hippocampal volume may explain a substantial part (almost half) of the detrimental effect of rs6859 (A) on AD risk. One potential explanation for this effect could be that *NECTIN2* is a key component of adherens junctions, playing a role in cell-cell adhesion and mediating the viral entry into the brain. Hence, its variation may affect the brain’s permeability and vulnerability to infections. Certain gene variants could lead to a greater burden of infection-induced brain damage, ultimately contributing to hippocampal atrophy. This hypothesis needs confirmation in further research.

Another notable finding is that the impact of the rs6859 polymorphism differed by hippocampal spheres. This seems likely as certain hippocampal regions are more susceptible than others in AD.^49^ The left and right hippocampus serve different cognitive functions, with the left associated with verbal memory and the right with spatial memory.^50^ Our results did not agree with a study that concluded no region-specific loss of the hippocampus in AD.^51^ However, the authors still found a greater atrophy of the hippocampus on the left side in semantic dementia cases. On the other hand, Lindberg and colleagues analysed the shapes of the hippocampus across multiple dementia subtypes and observed a consistent left hippocampal predominance in volume loss.^52^ Indeed, this aligns with our data, which includes participants with varying levels of cognition and those with AD.

It appears that a reduction in hippocampal volume due to an increase in the A allele of SNP rs6859 is likely to affect males only. In terms of statistical significance, a much-attenuated effect on right hippocampal volume was observed in males. Our findings stand in contrast to previous evidence from the UK Biobank. There, the results indicated that female carriers of the A allele of rs6859 with prior infections faced a risk of hippocampal loss, whereas males did not.^20^ Since, to our knowledge, no detailed studies have explored the gender-specific effects of *NECTIN2* on the brain or AD, we are unable to speculate on the underlying reasons. Our findings are novel but require replication in other cohorts.

AD is sometimes called type 3 diabetes due to certain commonalities with type 2 diabetes (T2D) pathology.^53^ Regardless, variable selection for the AD outcome did not support its inclusion in the final model. Published studies have often found conflicting results regarding the causal nature of the T2D-AD associations. Recently, what could be considered more reliable evidence from Mendelian randomization (MR) based on large GWAS has reported no direct connection between AD and T2D.^54^ That said, indirect pathways may still exist. For instance, our previous research using ADNI data demonstrated that diabetes negatively affects brain regions associated with AD.^18^ Diabetes strongly predicted hippocampal volume loss in our LMM models. We attribute this finding to the reported role of diabetes in causing abnormal activation of the hippocampus, which impairs learning and memory.^55^ Another plausible pathway is that diabetes can adversely impact hippocampal structure through disruptions in neurogenesis and neuroplasticity.^56^

Our mixed model analysis pointed to a relationship between frequent clinical visits and hippocampal atrophy, whereas in CMA, visits evinced a protective association. We infer that this discrepancy arises from the use of median visits in CMA, in contrast to MMA, where the relationship might be better captured with more granular data. The CMA primarily aims to identify a causal link between the A allele of rs6859 and AD counterfactually while treating visits as a covariate, which addresses a different research question altogether. Counterfactual mediation models are considered more capable than other mediation models, as they estimate real-world, interpretable effects, require fewer assumptions, and can be used to model exposure-mediator interaction effects. The smallest mediated effect was observed for the right hippocampal volume (RHV) at 42.75%, while the largest was found for the left hippocampal volume (LHV) at 49.76%. For the THV, the estimated effect was closer to that of the LHV, suggesting that the *NECTIN2* gene has a stronger impact on the left hippocampus and that the effect observed for the THV is largely due to the latter.

There are several possible mechanisms by which genetic variations in *NECTIN2* may affect hippocampal volume. *NECTIN2* is a relatively understudied gene, primarily due to its location near the popular *APOE4* and *TOMM40* locus.^12,57,58^ The SNP rs6859 is located in the non-coding region of the *NECTIN2* gene, and its polymorphisms may interfere with the binding of miRNAs, resulting in neurological damage.^59–61^ We checked the Human Protein Atlas to understand the impact of *NECTIN2*. While *NECTIN2* mRNA is present in the hippocampus, its expression has not been detected in either glial or neuronal cells, suggesting a potential post-transcriptional regulation or expression in other cell types.^62^

The *NECTIN2* gene is highly pleiotropic and influences multiple phenotypes, including a causal effect on LDL-C independent of the *APOE* effect.^63^ Elevation in LDL-C and other lipids has been shown to increase AD pathologic features markedly and is inversely correlated with hippocampal volume.^64,65^ The association between higher LDL-C and hippocampal volume was partially mediated by Aβ aggregation, underlining a complex web of pathological effects.^64^ Additionally, these *NECTIN2*-related changes could impact the health of astrocytes and neurons, which, in turn, may also affect the hippocampal mass.^66,67^ As described before, *NECTIN2* gene polymorphisms can also increase the synthesis of pTau.^30^

As regards the strengths of our study, the *medflex* estimate is robust in terms of outcome prevalence and non-collapsibility issues frequently encountered in the CMA of binary outcomes due to the use of counterfactual-based statistics.^68^ The robustness arises from how the counterfactual estimations are calculated, by estimating causal effects by simulating varying exposure-mediator relationships, which is quite distinct from computing conditional effects in traditional regression models. We also report significant improvement in understanding how the *NECTIN2* gene potentially influences AD risk through its endophenotypes, greatly increasing the explained variance to 69.16% from the previous 19.40%.^69^ Furthermore, we demonstrated statistically significant associations using longitudinal measures and summary data, systematically adjusting for established confounders.

Coming to the limitations, we were mainly constrained by the sample size, as hippocampal measurements were gathered for only a subset of participants in the cohort. This led to further reductions during data linkage. Other limitations include the lack of a racially diverse population in ADNI and the predominance of older individuals. Further studies are warranted to examine how the polymorphism affects the hippocampus in younger individuals.

Additionally, there may be confounders we have not accounted for, and a significant portion of the mediation effects linking the *NECTIN2* gene to AD remains unknown. Besides infections, weakened immunity related to aging and genetics can further accelerate neurodegeneration.^9^ We posit that studying inflammation in relation to the *NECTIN2* gene and AD may help explain part of the remaining unexplained pathway.

## Conclusions

This study found that hippocampal atrophy, reflected in lower hippocampal volume, is a significant mediator of the association between rs6859 (A) in the *NECTIN2* gene and increased AD risk in ADNI participants. Depending on the hippocampal region involved, this mechanism could account for nearly half the risk associated with rs6859 (A).

## Data Availability

All analyses in this study were conducted using publicly available datasets, which are accessible online.

https://adni.loni.usc.edu

## Abbreviations

AD: Alzheimer’s Disease
ADNI: Alzheimer’s Disease Neuroimaging Initiative
AIC: Akaike Information Criterion
CMA: Causal Mediation Analysis
HV: Hippocampal Volume
MRI: Magnetic Resonance Imaging
NDE: Natural Direct Effect
NIE: Natural Indirect Effect
ORQ: Ordered Quantile Normalization
SNP: Single Nucleotide Polymorphism
TE: Total Effect
T2D: Type 2 Diabetes
UTI: Urinary Tract Infections

## Funding

Research reported in this publication was supported by the National Institute on Aging of the National Institutes of Health under Award Numbers R01AG076019 and R01AG070487. The content is solely the responsibility of the authors and does not necessarily represent the official views of the National Institutes of Health.

## Acknowledgments

We would like to thank the ADNI team. Data collection and sharing for this project was funded by the Alzheimer’s Disease Neuroimaging Initiative (ADNI) (National Institutes of Health Grant U01 AG024904) and DOD ADNI (Department of Defense award number W81XWH-12-2-0012). ADNI is funded by the National Institute on Aging, the National Institute of Biomedical Imaging and Bioengineering, and through generous contributions from the following: AbbVie, Alzheimer’s Association; Alzheimer’s Drug Discovery Foundation; Araclon Biotech; BioClinica, Inc.; Biogen; Bristol-Myers Squibb Company; CereSpir, Inc.; Cogstate; Eisai Inc.; Elan Pharmaceuticals, Inc.; Eli Lilly and Company; EuroImmun; F. Hoffmann-La Roche Ltd and its affiliated company Genentech, Inc.; Fujirebio; GE Healthcare; IXICO Ltd.; Janssen Alzheimer Immunotherapy Research & Development, LLC.; Johnson & Johnson Pharmaceutical Research & Development LLC.; Lumosity; Lundbeck; Merck & Co., Inc.; Meso Scale Diagnostics, LLC.; NeuroRx Research; Neurotrack Technologies; Novartis Pharmaceuticals Corporation; Pfizer Inc.; Piramal Imaging; Servier; Takeda Pharmaceutical Company; and Transition Therapeutics. The Canadian Institutes of Health Research is providing funds to support ADNI clinical sites in Canada. Private sector contributions are facilitated by the Foundation for the National Institutes of Health (www.fnih.org). The grantee organization is the Northern California Institute for Research and Education, and the study is coordinated by the Alzheimer’s Therapeutic Research Institute at the University of Southern California. ADNI data are disseminated by the Laboratory for Neuro Imaging at the University of Southern California.

## Conflict of interest

The authors declare no conflict of interest.

## Contributions

S.U., K.G.A., and A.L.R. contributed to the study conception, data acquisition, data interpretation, and manuscript writing. O.B., and A.L.R. contributed to data analysis. K.G.A., S.U., and A.I.Y. contributed to data interpretation and manuscript revisions. All authors approved the final version of the manuscript.

## Ethics declarations

### Ethics approval and consent to participate

ADNI study methods were approved by institutional review boards of all participating institutions. The authors have only conducted secondary analyses of de-identified participant data. The ADNI obtained written informed consent for all ADNI participants by following local legislation and ADNI requirements. ADNI studies follow Good Clinical Practices guidelines, the Declaration of Helsinki, and United States regulations (U.S. 21 CFR Part 50 and Part 56).

## Consent for publication

Not applicable

## Supplementary Material

### Supplementary Figures

**Supplementary Figure S1. Study Flow Diagram**

**Supplementary Figure S2.** Histogram of hippocampal volume

**Supplementary Figure S3.** Smoothed trajectories of hippocampal volume with age, stratified by rs6859 allele status.

**Supplementary Figure S4.** Longitudinal change in hippocampal volume with clinical visits, stratified by rs6859 allele status

### Supplementary Tables

**Supplementary Table S1.** Male-specific regression estimates for the right hippocampal volume (n=165, observations=489)

**Supplementary Table S2.** Female-specific regression estimates for the right hippocampal volume (n=153, observations=413)

**Supplementary Table S3.** Male-specific regression estimates for the left hippocampal volume (n=165, observations=489)

**Supplementary Table S4.** Female-specific regression estimates for the left hippocampal volume (n=153, observations=413)

**Supplementary Table S5.** Multivariate regression estimates for rs6859 and covariates for THV in males

**Supplementary Table S6.** Multivariate regression estimates for rs6859 and covariates for THV in females

## Notes

### Competing Interest Statement

The authors have declared no competing interest.

