## Supplementary Materials for "Lower Hippocampal Volume Partly Mediates the Association Between rs6859 in the *NECTIN2* Gene and Alzheimer’s Disease: New Findings from Causal Mediation Analysis of ADNI Data"

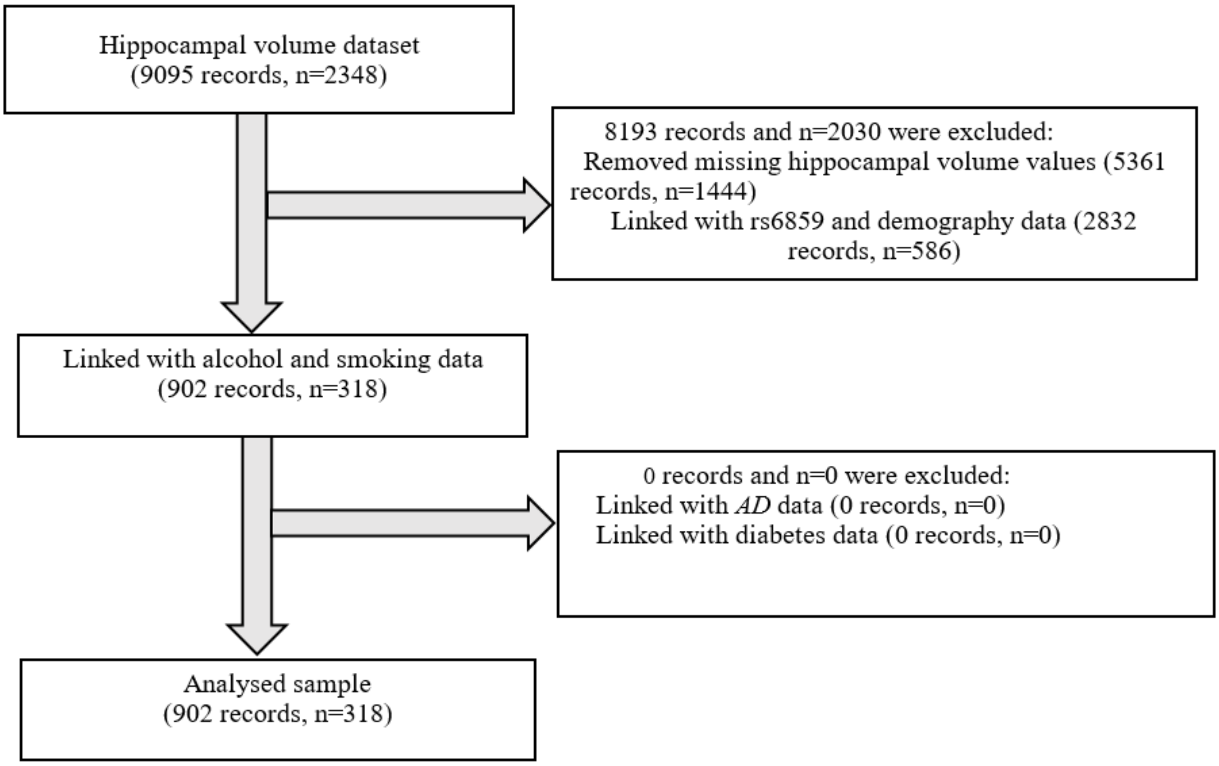


**Supplementary Figure S1.** Study Flow Diagram

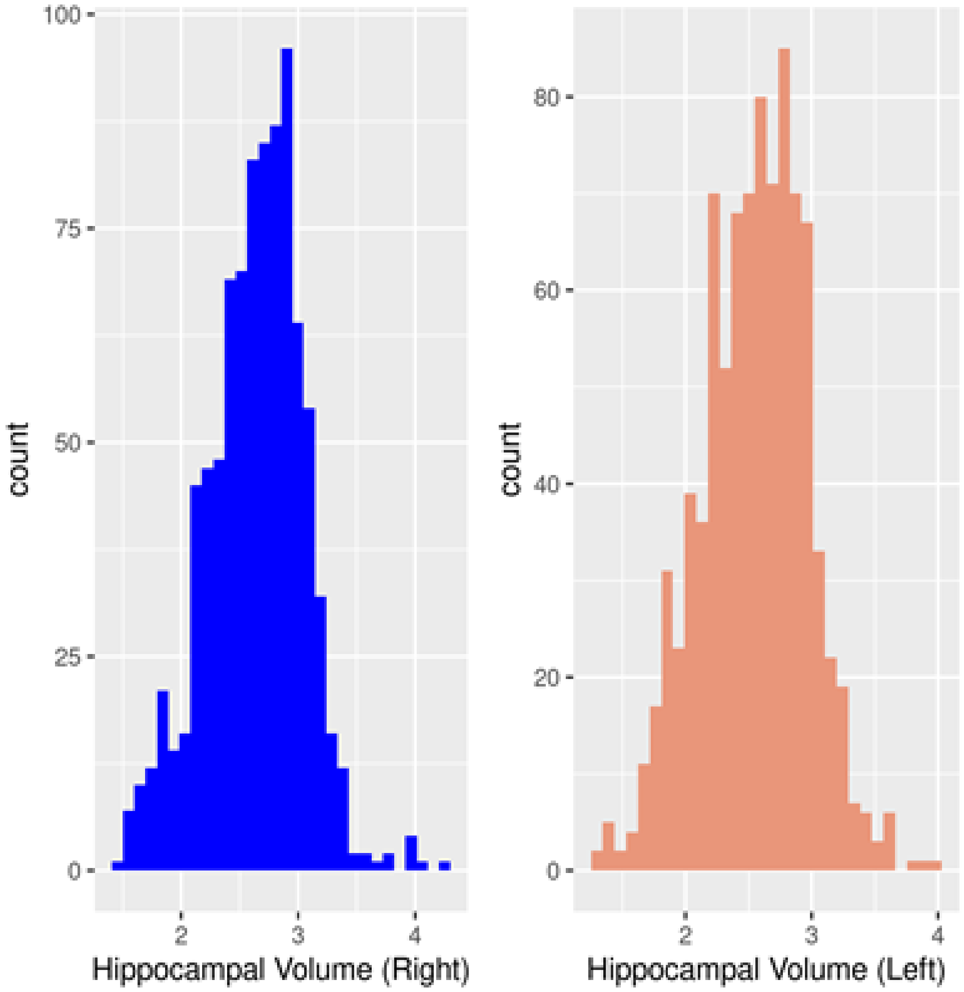


**Supplementary Figure S2.** Histogram of hippocampal volume

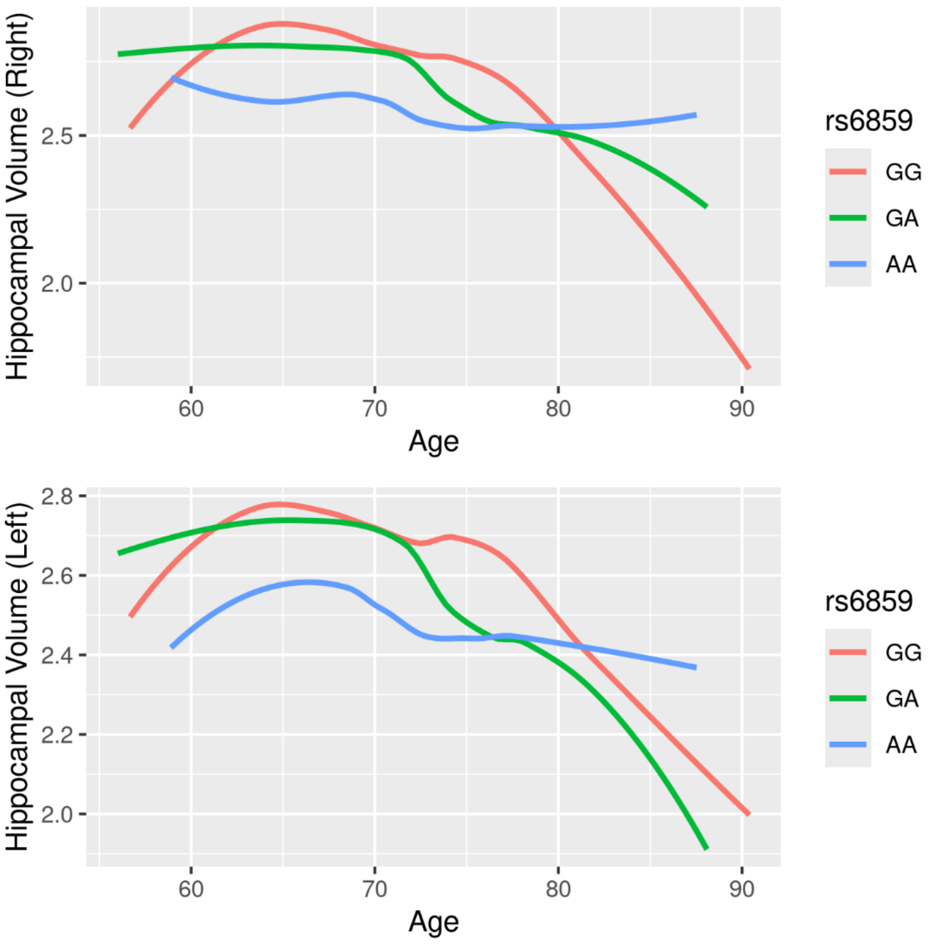


**Supplementary Figure S3.** Smoothed trajectories of hippocampal volume with age, stratified by rs6859 allele status.

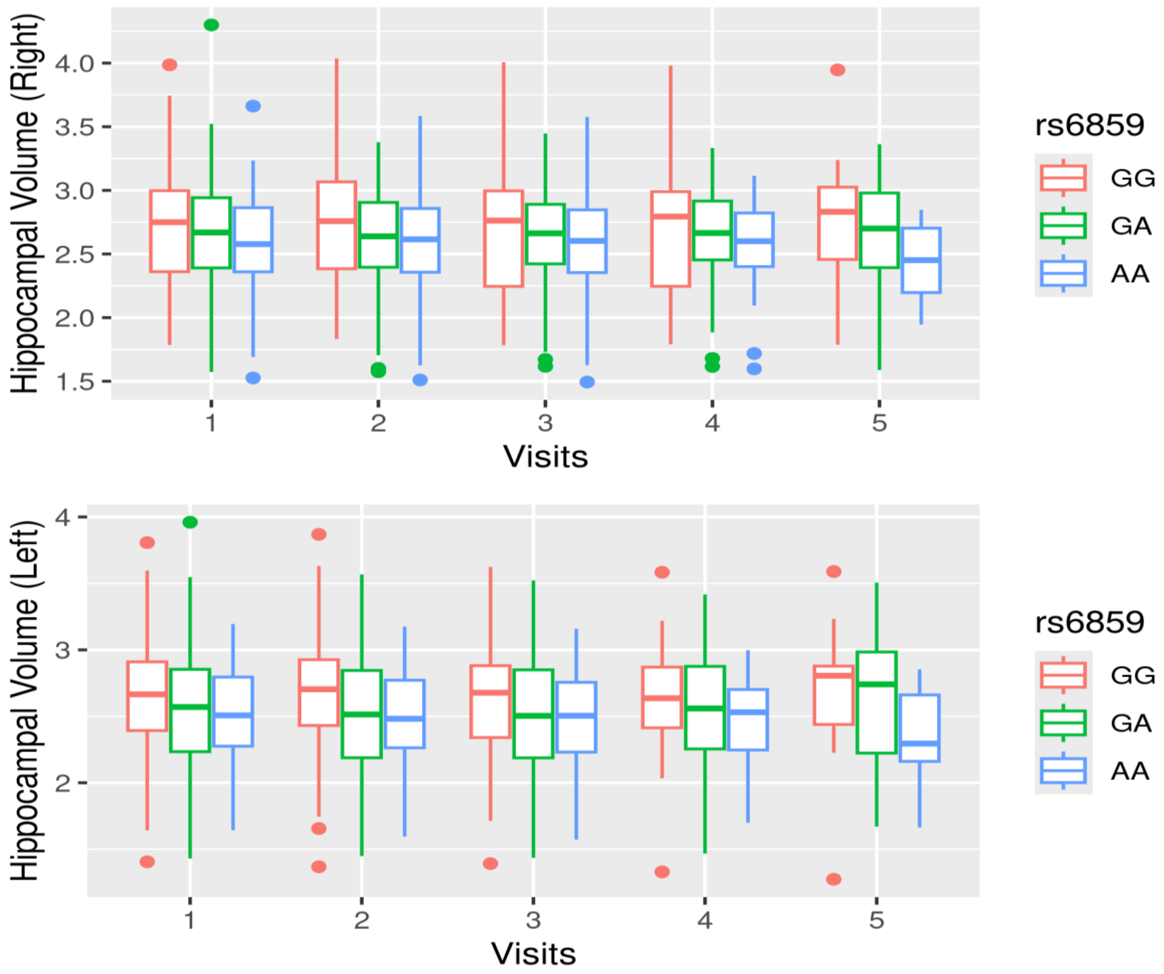


**Supplementary Figure S4.** Longitudinal change in hippocampal volume with clinical visits, stratified by rs6859 allele status

**Supplementary Table S1.** Male-specific regression estimates for the right hippocampal volume (n=165, observations=489)

| Variables | Estimates | 95% CI | *p* |
|---|---|---|---|
| Age (years) | -0.069 | -0.085, -0.053 | 0.000*** |
| Alcohol (ever) | -0.739 | -1.541, 0.061 | 0.070 |
| Diabetes (Yes) | -0.547 | -0.901, -0.192 | 0.002** |
| rs6859 | -0.181 | -0.374, 0.012 | 0.065 |
| Smoking (ever) | 0.308 | -0.014, 0.631 | 0.060 |
| Visits | -0.022 | -0.035, -0.009 | 0.000*** |

**Note.** The optimal set of variables in the model was selected using the AIC criterion by fitting a full model that included the following predictors: age, hippocampal volumes, diabetes status (yes/no), SNP rs6859, smoking status (ever/never), alcohol use (ever/never), number of visits, duration of education, race, and marital status (ever/never).

$^{*}p<0.05$; $^{**}p<0.01$; $^{***}p<0.001$.

**Supplementary Table S2.** Female-specific regression estimates for the right hippocampal volume (n=153, observations=413)

| Variables | Estimates | 95% CI | *p* |
|---|---|---|---|
| rs6859 | -0.094 | -0.296,0.107 | 0.365 |
| Age (years) | -0.056 | -0.073, -0.037 | 0.000* |
| Smoking (ever) | -0.181 | -0.566, 0.203 | 0.358 |
| Alcohol (ever) | 0.528 | -0.763, 1.820 | 0.429 |

| | | | |
|---|---|---|---|
| Visits | -0.022 | -0.039, -0.007 | 0.005** |
| Diabetes (yes) | -0.170 | -0.551, 0.211 | 0.388 |

**Note.** The optimal set of variables in the model was selected using the AIC criterion by fitting a full model that included the following predictors: age, hippocampal volumes, diabetes status (yes/no), SNP rs6859, smoking status (ever/never), alcohol use (ever/never), number of visits, duration of education, race, and marital status (ever/never).

$^{*}p<0.05$; $^{**}p<0.01$; $^{***}p<0.001$.

**Supplementary Table S3.** Male-specific regression estimates for the left hippocampal volume (n=165, observations=489)

| **Variables** | **Estimates** | **95% CI** | ***p*** |
|---|---|---|---|
| Age (years) | -0.063 | -0.079, -0.048 | 0.000*** |
| Diabetes (yes) | -0.484 | -0.829, -0.139 | 0.006** |
| rs6859 | -0.242 | -0.435, -0.050 | 0.014* |
| Visits | -0.022 | -0.035, -0.010 | 0.000*** |

**Note.** The optimal set of variables in the model was selected using the AIC criterion by fitting a full model that included the following predictors: age, hippocampal volumes, diabetes status (yes/no), SNP rs6859, smoking status (ever/never), alcohol use (ever/never), number of visits, duration of education, race, and marital status (ever/never).

$^{*}p<0.05$; $^{**}p<0.01$; $^{***}p<0.001$.

**Supplementary Table S4.** Female-specific regression estimates for the left hippocampal volume (n=153, observations=413)

| **Variables** | **Estimates** | **95% CI** | ***p*** |
|---|---|---|---|
| Age (years) | -0.053 | -0.069, -0.036 | 0.000*** |
| Education (years) | 0.048 | -0.000, 0.097 | 0.057 |
| Married Status (never married) | 0.486 | 0.005, 0.968 | 0.051 |
| Race (White) | -0.357 | -0.815, 0.100 | 0.131 |
| Visits | -0.027 | -0.042, -0.013 | 0.000*** |

**Note.** The optimal set of variables in the model was selected using the AIC criterion by fitting a full model that included the following predictors: age, hippocampal volumes, diabetes status (yes/no), SNP rs6859, smoking status (ever/never), alcohol use (ever/never), number of visits, duration of education, race, and marital status (ever/never).

$^{*}p<0.05$; $^{**}p<0.01$; $^{***}p<0.001$.

**Supplementary Table S5.** Multivariate regression estimates for rs6859 and covariates for THV in males

| **Variables** | **Estimates** | **95% CI** | ***p*** |
|---|---|---|---|
| Age (years) | -0.071 | -0.086, -0.056 | <0.001*** |
| Diabetes (yes) | -0.550 | -0.898, -0.202 | 0.002** |
| rs6859 | -0.216 | -0.407, -0.026 | 0.028* |

| | | | |
|---|---|---|---|
| Visits | -0.021 | -0.033, -0.009 | 0.000*** |
| Smoking (ever) | 0.242 | -0.073, 0.559 | 0.139 |
| Alcohol (ever) | -0.655 | -1.442, 0.132 | 0.108 |

**Note.** The optimal set of variables in the model was selected using the AIC criterion by fitting a full model that included the following predictors: age, hippocampal volumes, diabetes status (yes/no), SNP rs6859, smoking status (ever/never), alcohol use (ever/never), number of visits, duration of education, race, and marital status (ever/never).

$^{*}p<0.05$; $^{**}p<0.01$; $^{***}p<0.001$.

**Supplementary Table S6.** Multivariate regression estimates for rs6859 and covariates for THV in females

| **Variables** | **Estimates** | **95% CI** | ***p*** |
|---|---|---|---|
| Age (years) | -0.058 | -0.075, -0.042 | 0.000*** |
| Visits | -0.022 | -0.038, -0.007 | 0.003** |
| Education (years) | 0.046 | -0.005, 0.098 | 0.079 |
| Marriage (never married) | 0.530 | 0.020, 1.041 | 0.042* |

**Note.** The optimal set of variables in the model was selected using the AIC criterion by fitting a full model that included the following predictors: age, hippocampal volumes, diabetes status (yes/no), SNP rs6859, smoking status (ever/never), alcohol use (ever/never), number of visits, duration of education, race, and marital status (ever married/never married).

$^{*}p<0.05$; $^{**}p<0.01$; $^{***}p<0.001$
